# Assessing Parent-cocreated Sensory Reactivity Outcomes in Children with Neurodevelopmental Disorders Undergoing Bumetanide Treatment: A Multiple-Baseline Single-Case Experimental Design

**DOI:** 10.64898/2026.04.22.26351464

**Authors:** L.M.G. Geertjens, G. Cristian, J. R. Ramautar, L. Haverman, B.D. Schalet, K. Linkenkaer-Hansen, G.J van der Wilt, J.J Sprengers, H. Bruining

**Affiliations:** Amsterdam UMC, N=You Neurodevelopmental Precision Center, Department of Child and Adolescent Psychiatry & Psychosocial Care, Amsterdam, the Netherlands; IQ Health, Radboud University Medical Center, Nijmegen, The Netherlands; Amsterdam UMC, Emma Center for Personalized Medicine, Amsterdam, the Netherlands; Amsterdam Public Health Research Institute, Methodology and Mental Health and Personalized Medicine, Amsterdam, The Netherlands; Amsterdam UMC, Emma Children’s Hospital, Department of Child and Adolescent Psychiatry & Psychosocial Care, Amsterdam, the Netherlands; Department of Epidemiology and Data Science, Amsterdam UMC, Amsterdam, the Netherlands; Department of Integrative Neurophysiology, Center for Neurogenomics and Cognitive Research (CNCR), Vrije Universiteit Amsterdam, The Netherlands

**Author notes:** **Correspondence to** Hilgo Bruining, MD, PhD, Department of Child and Adolescent Psychiatry & Psychosocial care, Meibergdreef 9, 1105 AZ, Amsterdam, Netherlands. shared author positions (L.M.G. Geertjens, G. Cristian, J.J Sprengers, H. Bruining).

**Keywords:** neurodevelopmental disorder, single case experimental design (SCED), patient reported outcome measures (PROMs), bumetanide

## Abstract

Progress in pharmacological treatment development for neurodevelopmental disorders is hindered by a misalignment between targeted mechanisms, outcome measures, and trial designs. This study was initiated as a post-trial access pathway for bumetanide and later expanded with treatment-naive participants. Within this framework, we implemented a parent-cocreated sensory outcome measure set (PROMset) in an unmasked, multiple-baseline single-case experimental design with randomized baseline periods of 2–12 weeks, followed by 6 months of bumetanide treatment (up to 1.5 mg twice daily). Participants (7–19 years) had atypical sensory reactivity and a diagnosis of ASD, ADHD, epilepsy, or TSC.

The primary outcome was a PROMset comprising seven PROMIS item banks assessing anxiety, depressive symptoms, sleep disturbance, fatigue, sleep-related impairment, cognitive function, and peer relationships. Secondary outcomes included SSP, SRS-2, RBS-R, and ABC. Of 113 enrolled participants (mean age 13.2 [SD 2.7], 64% male), 102 completed the trial and 95 had analyzable PROMsets. At baseline, PROMset scores showed substantial impairment across domains (mean deviation ≥9.0 T-score points, p<.001) and correlated with sensory reactivity (SSP; r≥-0.40, p<.001). Individual-level analyses showed improvement in 24–41% of participants per PROM domain, most frequently in anxiety and depressive symptoms (41% and 38%; mean across-case Cohen’s d≈-1). Overall, 83% improved on at least one domain. Group-level analyses showed improvement across all secondary outcomes (p<.001), with superiority over historic placebo for RBS-R and SSP.

Integrating PROMsets with individualized trial designs can reveal clinically meaningful changes, supporting a more sensitive and patient-centered framework for treatment evaluation in heterogeneous populations.

## Introduction

Several promising drug repurposing strategies have recently been explored across neurodevelopmental disorders (NDDs), including Autism Spectrum Disorder (ASD) and intellectual disabilities (Baribeau, Vorstman, & Anagnostou, 2022; Cortese et al., 2024). For example, arbaclofen, balovaptan, bumetanide, and oxytocin showed potential in preclinical studies on genetic animal models (Ben-Ari, 2017; Henderson et al., 2012; Qin et al., 2015; Sala et al., 2011). However, validation in human trials has proven difficult and none of these treatments has been approved as on-label medication for NDDs (Daniels et al., 2023; Fuentes et al., 2023; Hollander et al., 2022; Jacob et al., 2022; Sikich et al., 2021; Veenstra-VanderWeele et al., 2017).

One key challenge is the lack of outcome measures sufficiently sensitive to detect meaningful clinical change (Brugha, Doos, Tempier, Einfeld, & Howlin, 2015). Many studies rely on diagnostic instruments designed for classification rather than change, which lack the psychometric sensitivity to detect treatment-related effects over time. At the same time, there is growing recognition of the importance of evaluating interventions from the patient perspective, focusing directly on how individuals feel, function, and navigate daily life (Crossnohere et al., 2025). Together, these considerations hightlight the need for more refined outcome measures, that capture everyday functioning and extend beyond short-term symptom change to include repeated assessments of real-world functioning and longer-term developmental trajectories.

Daily functioning in children with NDDs is strongly shaped by how they adapt and respond to sensory stimuli (Gentil-Gutierrez, Cuesta-Gomez, Rodriguez-Fernandez, & Gonzalez-Bernal, 2021; Mulligan, Douglas, & Armstrong, 2021). Sensory processing difficulties can directly interfere with participation in everyday activities, learning, and social interaction, making sensory reactivity a clinically meaningful treatment target. Accordingly, atypical sensory reactivity is included as a core diagnostic feature of ASD (*Diagnostic and statistical manual of mental disorders*, 2013). Despite this recognition, sensory-related outcomes are rarely included in intervention studies, largely due to the limited availability of sensitive assessment tools and the inherently variable and context-dependent nature of sensory reactivity responses.

To address this need, we previously conducted interviews and focus groups with parents of children with ASD to identify relevant sensory reactivity-related outcomes (van Andel et al., 2021). This resulted in the identification of 11 health-related outcomes, including behavior that is directly motivated by sensory reactivity as well as daily life outcomes that are indirectly affected. Direct relevant outcomes included sensory stimulation tolerance and sensitivity to sensory stimuli. Indirect relevant outcomes involved irritable behavior, anxiety, mood problems, sleep problems, fatigue, physical complaints, daily functioning and participation, routines, structures and dealing with change, and social interaction and communication (van Andel et al., 2021). Based on these insights and FDA guidance, we developed a parent-proxy patient-reported outcome measure set (PROMset) using existing, validated item banks from the Patient-Reported Outcomes Measurement Information System (PROMIS®). PROMIS® comprises standardized questionnaires designed to assess physical, mental, and social health from the patient’s perspective (Cella et al., 2010). PROMIS® uses modern psychometric methods, including Item Response Theory and computer-adaptive testing, to generate precise and comparable scores across conditions. Most of the PROMs in the PROMset have been psychometrically validated in Dutch populations, complementing their prior validation in American cohorts (Westerweel et al., 2026). Together, these features support the PROMset as a comprehensive and accessible composite outcome measure for use in clinical trials (van Andel et al., 2021).

Alongside the need for more sensitive measures, there is a parallel requirement for trial designs to capture individual variability. Traditional randomized controlled trials (RCTs) prioritize group-level mean differences, potentially obscuring clinically meaningful effects in heterogeneous NDD populations (Beversdorf et al., 2022; McCracken et al., 2021; Sprengers, Geertjens, & Bruining, 2024). Trials evaluating bumetanide, a diuretic drug being repurposed for ASD on the basis of its actions on GABAergic signaling, illustrate this issue: While Phase-II trials reported promising improvements on social communication (Lemonnier et al., 2012; Lemonnier et al., 2017; Zhang et al., 2020), repetitive behaviors (Sprengers et al., 2021) and irritability (van Andel et al., 2022; van Andel et al., 2020), they also revealed significant inter-individual variation. This variability likely contributed to the premature termination of a Phase-III trial, where changes on traditional outcomes failed to exceed the placebo effect (Fuentes et al., 2023). These findings highlights a potential “signal-loss” inherent in group-level designs. Single-case experimental designs (SCEDs) offer an alternative approach by using frequent, within-subject measurements, treating variability as informative rather than noise. This approach enables detection of individualized treatment trajectories that may be missed in traditional RCTs. These individual-level evaluations can help identify responsive subgroups, moving beyond a one-size-fits-all framework toward more targeted intervention strategies.

In our previous bumetanide trials, we observed improvement of symptons that were not readily captured by the conventional outcome scales, such as changes in energy, fatigue and social participation; symptoms that indeed were also indentified in the PROMset mentioned above. Motivated by these observations, the present study was intiatied as a post-trial access pathway for bumetanide and designed as a proof-of-concept evaluation of two methodological innovations: the use of a set of parent-cocreated PROMs measuring daily functioning related to sensory reactivity and the use of SCEDs to evaluate individual trajectories. During ethical review, the committee advised expansion of the study to include a treatment-naïve cohort, in order to strengthen interpretability and reduce potential bias. Consequently, the study comprised both participants with prior bumetanide exposure and newly enrolled, treatment-naïve individuals. Using repeated, within-subject assessments, we examined whether the PROMset could capture clinically meaningful sensory-related and functional changes over time in children and adolescents with NDDs receiving bumetanide. Rather than establishing definitive efficacy, the study aimed to (i) evaluate the feasibility and sensitivity of the PROMset, (ii) characterize individual-level response patterns using SCEDs, and (iii) explore how these outcomes relate to conventional group-level measures. In doing so, this study provides empirical insight into whether alternative outcome strategies can detect treatment-relevant changes that may be missed by traditional trial designs.

## Methods

### Study design and participants

The study protocol was previously published (Geertjens et al., 2022); here we summarize key elements and deviations. The Post-Trial Access Cohort Bumetanide for Developmental Disorders (BUDDI) trial was an open-label, single-center, non-masked, multiple-baseline SCED at the N=You Center for NDD at Amsterdam UMC, a nationwide tertiary out-patient department in the Netherlands. A multiple-baseline SCED was preferred over n-of-1 or placebo-run-in designs due to ethical concerns regarding reallocation to placebo in prior trial participants and the risk of carry-over effects of bumetanide (Sprengers et al., 2021; van Andel et al., 2022). In addition, a phase-III trial addressing placebo effects was ongoing at study initiation (Fuentes et al., 2023). On advice of the ethical review board, enrollment was expanded to include treatment-naïve participants, to strengthen interpretability and accommodate community interest.

Eligible participants were individuals who 1) participated in previous bumetanide trials (Sprengers et al., 2021; van Andel et al., 2022; van Andel et al., 2020) or 2) new participants aged 7-18 years meeting prior inclusion criteria: a) autism diagnosis on DSM-IV/V and Social Responsiveness Scale t-score ≥60; b) expert-confirmed diagnosis of ASD, ADHD, or epilepsy and a clinical score (+/-1SD) on the short sensory profile; c) definite diagnosis of Tuberous Sclerosis Complex with history of behavioral problems. Exclusion criteria included: body weight <30 kg; concomitant use of methylphenidate or other psychostimulants; start of behavioral therapy or unstable dose of concomitant psychoactive medication, two months prior to study participation; chronic renal disease; unstable serious illness; use of nonsteroidal anti-inflammatory drugs; documented history of hypersensitivity reaction to sulphonamide derivatives and/or inability to comply with the study and safety procedures.

The study adhered to the Declaration of Helsinki and Good Clinical Practice. The protocol was approved by the medical ethical review board of UMC Utrecht. Study safety was overseen by the monitoring board of Amsterdam. All participants or their legal representatives signed informed consent and received no financial compensation. This study was registered (EudraCT 2020–002196-35) and followed CONSORT-CENT guidelines.

### Randomization and masking

Participants were randomized to baseline durations of 2-12 weeks in one-week intervals, ensuring equal distribution via a SAS-generated list. During the second visit, participants were assigned to the next open slot on the list; neither researchers nor participants were masked for baseline duration.

### Procedures

During the first visit, participants underwent medical screening (including weight, blood pressure, and cardiac family history), clinical history taking and EEG. When available, historic IQ tests results were obtained, for which the most recent reliable result was carried forward. Caregivers were instructed to activate an account in the KLIK portal for online administration of the PROMset and conventional instruments (van Muilekom et al., 2022). The second visit included cognitive testing and laboratory evaluations.

During the 6-month intervention, participants received oral bumetanide tablets twice-daily, starting with 2dd0.5mg. At day 7 (D7), the dose was increased to 2dd1.0mg if tolerated. In individuals with a body weight >45kg, the dose could be increased to 2dd1.5mg at D28. Safety monitoring followed prior protocols (Sprengers et al., 2021; van Andel et al., 2022; van Andel et al., 2020), and consisted of physical examination to check for dehydration or hypotension at D14, D28, D56, D91, D119, D154, and D182 and laboratory evaluations to check for dehydration and hypokalemia at D14, D28, D56 and D91. All participants received oral supplementation with 0.5mmol/kg potassium chloride to prevent hypokalemia from the start of treatment.

The primary outcome (PROMset) was assessed weekly; secondary outcomes at baseline, D91 and D182. EEGs were obtained at baseline, before the first administration of bumetanide, 1.5 hours after first administration (Tmax), D28, D56, D91, D119, D154, and D182.

Neurocognitive outcomes were evaluated at baseline, D91 and D182. The results of EEG and neurocognitive outcome analyses will be published separately.

### Outcomes

The primary outcome included the PROMIS item banks (proxy version) of the aforementioned PROMset, three item banks had to be excluded because of delays in the translation to Dutch (see protocol (Geertjens et al., 2022)). Computer adaptive testing (CAT) was used where possible, otherwise PROMIS short forms (SF) were used. CAT makes use of item response theory to select the PROM items most suited for defining the estimated severity score, significantly limiting the number of questions asked and thereby decreasing the burden for parents (Luijten et al., 2020) (Time burden decreases from 8-10min to 1-2min per PROMIS item bank). The PROMIS parent proxy measures included were: SFv2.0 – Anxiety 8a(Irwin et al., 2012), SFv1.0 - Cognitive Function 7a, CATv2.0 - Depressive Symptoms, CATv1.0 - Sleep Disturbance, CATv2.0 – Fatigue, CATv1.0 - Sleep-Related Impairment and CATv2.0 - Peer Relationships (Irwin et al., 2012). PROMIS scores are reported using a T-score metric, that ranges from 20 to 80, with general population means for these versions falling between 40 and 50, with a standard deviation of 10 (Carle, Bevans, Tucker, & Forrest, 2021). Higher scores indicate that the measured construct is more present. To maintain the scientific rigor of the weekly SCED measurements while minimizing participant burden, other questionnaires from the broader PROMset were transitioned to secondary outcomes, ensuring that high-frequency data collection remained feasible for families over the 6-month period.

Secondary outcomes were measured three-monthly and included all conventional questionnaires implemented in our previous bumetanide trials (Sprengers et al., 2021; van Andel et al., 2022; van Andel et al., 2020): Social Responsiveness Scale-2 total score (SRS-2; range 0‐195; higher score indicates more affected), Repetitive Behavior Scale‐Revised (RBS-R; range 0‐129; higher score indicates more affected), Short Sensory Profile (SSP; range 38‐190, lower score is more affected), Aberrant Behavior Checklist (ABC) irritability subscale (ABC-I; range 0‐45, higher score is more affected), hyperactivity subscale (ABC-H; range 0-48; higher score is more affected), and lethargy subscale (ABC-L; range 0-48; higher score is more affected), Parent Global Impression – severity scale (PGI-S; range 0-7, higher is more affected), Parent Global Impression – improvement (PGI-I; range 0-7, 4 is no improvement, lower is improving, higher is worsening). All submitted questionnaires were complete.

The combination of PROMIS item banks and conventional questionnaires allows us to evaluate all themes elicited in our sensory reactivity PROM identification study (van Andel et al., 2021), except for ‘physical stress experience’.

Adverse events were documented according to severity, duration, attribution, and outcome with the National Cancer Institute Common Terminology Criteria for Adverse Events rating scale and classified in Medical Dictionary for Regulatory Activities categories.

### Statistical analysis

Power calculations were based on group effect size obtained from our previous trial (BAMBI), which reported a Cohen’s d of 0.37 (80% Confidence Interval: 0.23–0.53). With a sample size of at least 60 participants, the power to detect an aggregated across case effect size of d=0.4 was calculated to be 0.69, even under conditions of low to medium autocorrelation of the outcome measures (r=0.3) (Bouwmeester & Jongerling, 2020). We anticipated that approximately 75 individuals from prior cohorts would participate in the current study, including a minimum of 20 participants who had been previously allocated to the placebo group. To achieve a total sample size of 60 treatment-naïve individuals, we aimed to include at least 40 new participants in the current cohort. Consequently, the total intended sample size for this study was set at 115

Baseline PROMset results were compared with reference populations using t-tests or Wilcoxon signed rank test, depending on the distribution of the data. Data from Carle et.al (2021) was used as the primary reference (Carle et al., 2021). This dataset did not include cognitive function for which the standard PROMIS American population was used (HealthMeasures, 2025). Pearson correlation coefficients were used to examine the association between baseline PROMset scores and SSP scores. Outcomes were analyzed for the per protocol population.

Visual inspection is a commonly used analytical method in SCEDs (“Visual analysis of single-case intervention research: Conceptual and methodological issues,” 2014), and its rigor can be improved using visual aids like the 2-standard deviation (2SD) band alongside formal statistical tests. This study employed two such tests: Interrupted Time Series (ITS) analysis and randomization tests. ITS accounts for baseline instability and estimates changes in mean and trajectory during treatment, while randomization tests avoid assumptions such as normality and random sampling, which are often violated in SCEDs. While statistical testing using several methods validates visual analysis results, marked visual effects rule out false positive statistical results, allowing us to validate the results more robustly and thereby enhancing the replicability of findings in real-world settings. Analyses were conducted at the individual level and across-cases to assess treatment effects in the full sample. Individual response was defined as improvement on visual-2SD-inspection and a significant result on one or both other formal statistical testing strategies (Kratochwill & Shadish, 2010). By integrating these analytical strategies, we aimed to provide a comprehensive evaluation of the treatment’s impact on sensory reactivity across the participant cohort. More detailed information on the tests of Visual-2D-Inspection, ITS and randomization test is provided in the supplementary methods (see appendix S1; Table S1).

Secondary outcomes were analyzed with linear models including baseline measurement, age, and sex to correct for confounding factors. Residual normality assumptions were tested by residual plots. Mean differences after treatment were calculated with 95%CI and p-values. Treatment interactions with sex and age were tested with likelihood ratio tests. In an attempt to mitigate the risk of false positive findings and leverage the structural advantages of the post-trial access design, linear mixed models were used as described earlier (Sprengers et al., 2021) to compare treatment effects to historic placebo conditions at three months. Pearson correlation coefficients were used to examine the association between changes in PROMset scores and changes in SSP scores.

The list for baseline duration was generated in SAS v9.4M6 (SAS Institute, Cary, NC), all other analyses were performed with RStudio 2023.12.1+402 “Ocean Storm” Release.

## Results

### Participant Characteristics

Participants were enrolled between December 9 2020 and March 22 2023 when the target sample size was exceeded. Of 109 prior trial participants approached, 93 received study information, and 32 consented and were enrolled. Among new participants, 690 caregivers spontaneously expressed interest following media coverage of the previous trials. Of these, 187 caregivers received study information (first-come-first-served), 87 consented and were enrolled, and 86 were notified that the intended inclusion number was reached. Thus, in total 119 participants were allocated to baseline. Six participants did not advance to intervention because of an inability to adhere to study protocol (n=5), or withdrawal of consent (n=1). Finally, 113 participants started the intervention (Figure 1). Baseline characteristics (Table 1) did not differ between those who did and did not proceed (p≥0.09).

**Table 1:**
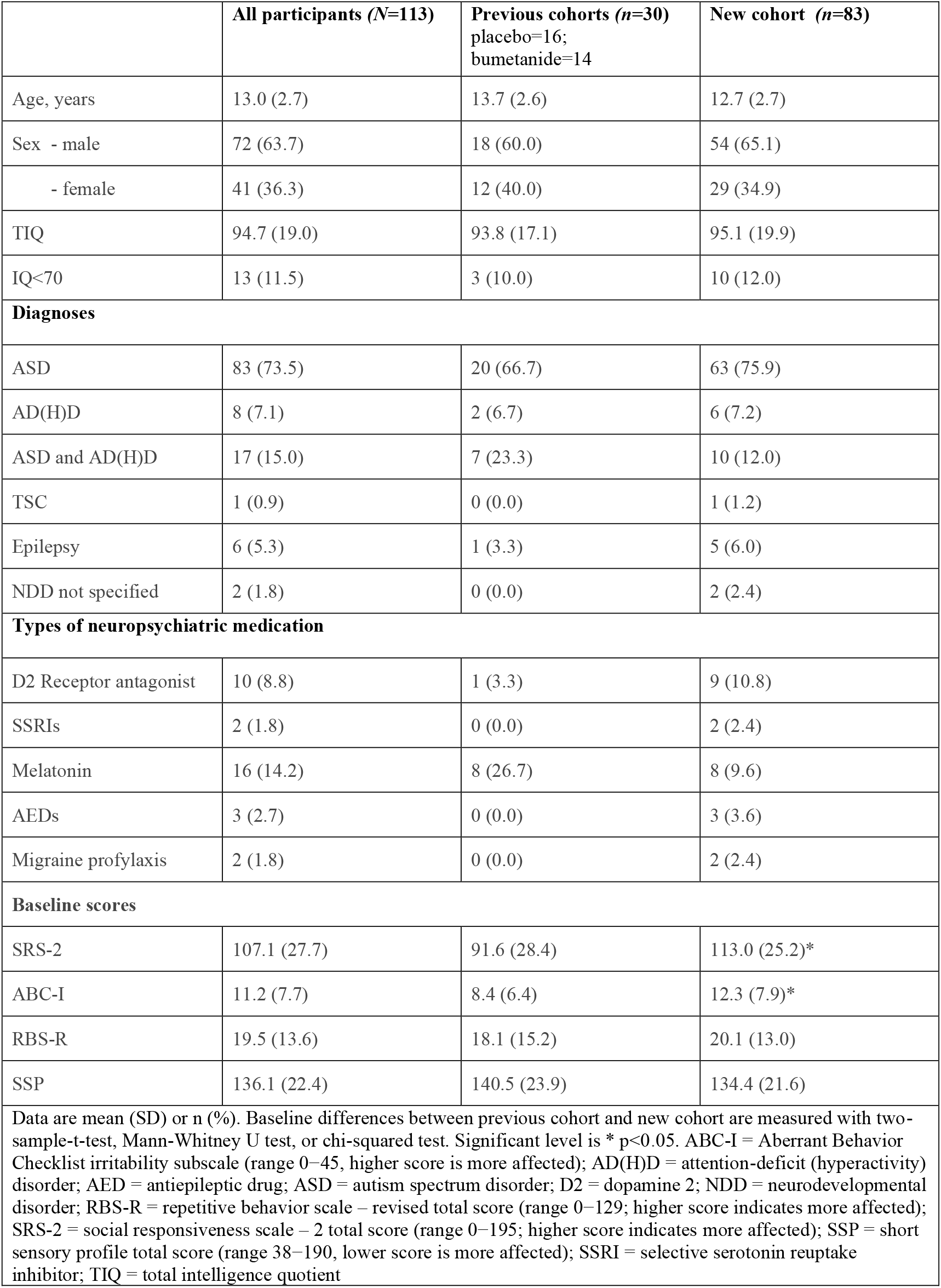
Baseline characteristics of the intention-to-treat population.

**Figure 1:**
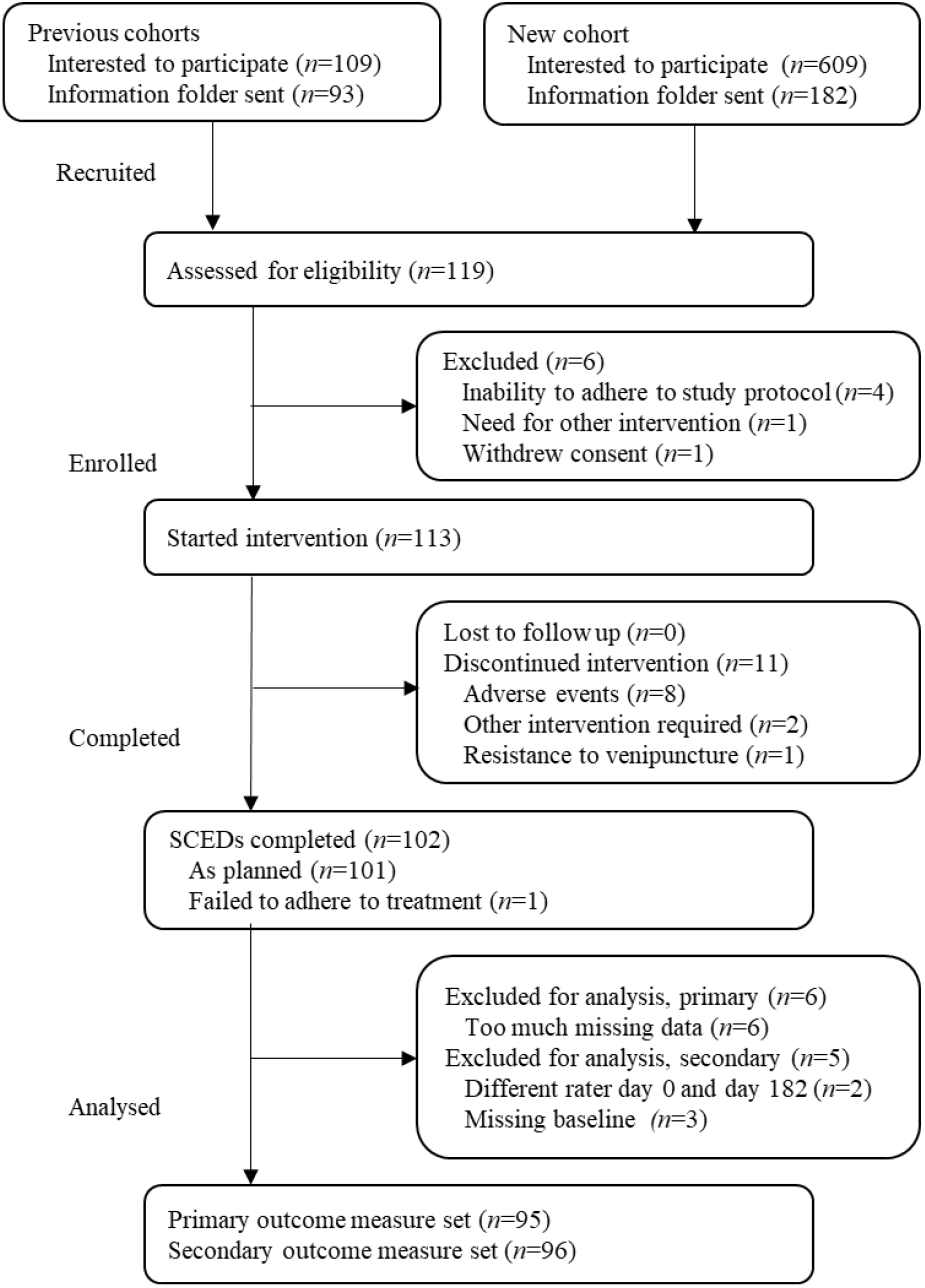
Trial profile of the study

Of the 113 participants who began the intervention, 101 completed the trial as planned. One completed the trial, without adhering to treatment. Nine discontinued due to adverse events: one experienced decreasing kidney function, while eight had diuretic events with mild hypokalemia. Additionally, one participant withdrew due to difficulties with venipunctures, and two stopped because they required other behavioral therapy. For the primary outcome analysis, six participants were excluded due to insufficient baseline data, meaning they did not have enough measurements to assess baseline variability; five of them were randomized to baseline periods of 2-4 weeks. In terms of secondary outcomes, five participants were excluded from analysis due to missing data (Figure 1).

Treatment adherence was monitored through interviews, drug diary, and inspection of returned medication strips. The mean bumetanide dose was 0.04mg/kg/day (range 0.01 to 0.07mg/kg/day). At D7, the dose was increased to 1.0mg twice-daily for 98 participants, while the remaining five participants had their doses increased at a later time. 67 participants were eligible for a dose increase to 1.5mg twice-daily, and this increase was implemented for 60 of them. In 12 cases, the dose remained below the target due to nonspecific somatic complaints (*n*=10) and persistent hypokalemia (*n*=2). Additionally, the dose was temporarily reduced for 12 other participants because of hypokalemia.

### Baseline PROMIS scores

Participants showed significant deviations from normative PROMIS values across all domains (Mean difference [MD] |9.0-14.6|, p<0.001; Figure 2B; Table S2). The largest differences were observed for fatigue (MD=14.6), anxiety (MD=11.7), and cognitive function (MD=‐11.6).

**Figure 2:**
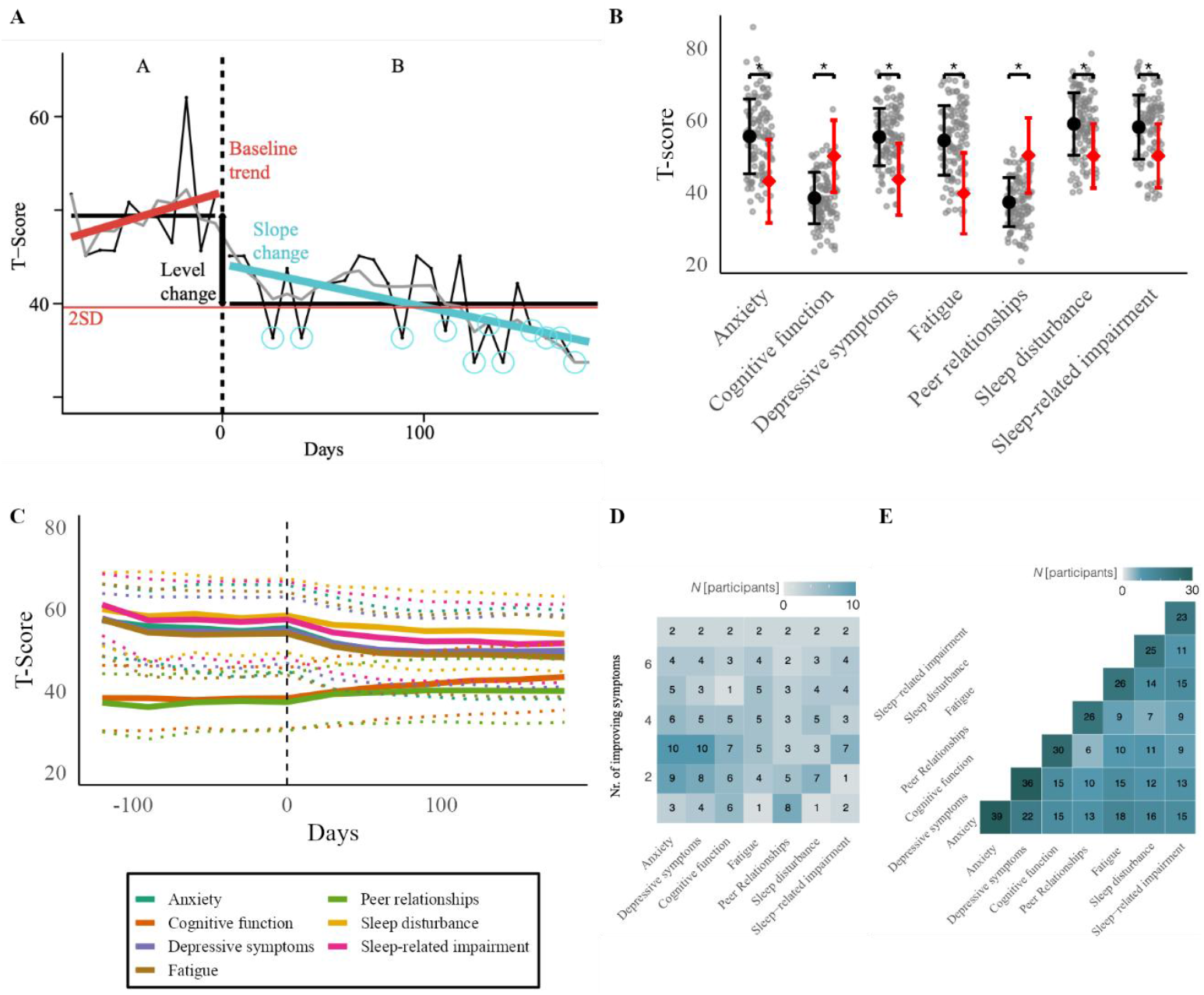
Primary and secondary outcomes of the study. (A) Visualization of the statistical methods used to analyze treatment effects on the PROMIS item banks. Shown is a longitudinal mapping of the item bank scores. The vertical dotted line indicates start of intervention (D0), which separates the randomized baseline phase (A) from the intervention phase (B). The thick black horizontal lines indicate the mean PROM item bank scores per phase. For the visual-2SD method a 2SD band is graphed (thin red horizontal line). Observations outside of the 2SD band in the intervention phase are marked with a blue circle. Two consecutive observations outside the 2SD band is labelled as a significant change. The interrupted time series analyses significant level and slope changes. Level change (thick black vertical line) is the difference in mean baseline and mean intervention score. Slope change is the difference in baseline slope(thick red line) and intervention slope (thick blue line). (B) Visualization of baseline PROMIS item bank scores. Grey dots represent t-scores of each participant. Sample mean ±1SD are shown in black dot and error bars and reference population mean ±1 SD are shown in red. Group differences were tested with one sample t-test or Wilcoxon signed rank test. (C) Visualization of aggregated PROMIS item bank scores over time. Shown are mean ±1SD PROM scores over time. The vertical dotted line indicates start of intervention (D0). (D) Heatmap depicting how many participants respond on multiple PROMs. (E) Heatmap showing the combination of PROMIS item bank responders to show whether response in specific measures tend to cluster together. PROM= patient reported outcome measure (higher scores reflect poorer functioning except for cognitive function and peer relations where higher scores reflect better functioning); Significance level * p<0.05.

SSP scores correlated moderately with PROMIS measures of anxiety, cognitive function, depressive symptoms, and fatigue (Pearson *r* ≈ 0.4), supporting a meaningful association between sensory reactivity and a range of mental and physical health domains captured by the PROMset (Figure S1).

### Individual effects

A central aim of this study was to demonstrate the ability of the SCED framework to detect individual-level treatment effects in addition to across-cage average effects. Consistent with this aim, analyses using interrupted time series and randomization tests showed significant improvements across most PROMset domains (*p*<.001). However, individual-level analyses revealed substantial heterogeneity in treatment response and provided greater clinical resolution. Across PROM domains, 24–41% of participants showed statistically significant improvement at the individual level (Table 2; Figure 2C; Figure S2; Table S3). Among those showing improvement, effect sizes were frequently large, with Cohen’s *d* exceeding ±1 in 34– 56% of individuals across PROMs. Overall, 83% of participants improved on at least one PROM, 57% improved on two domains, and 36% improved on three domains (Figure 2D).

**Table 2:**
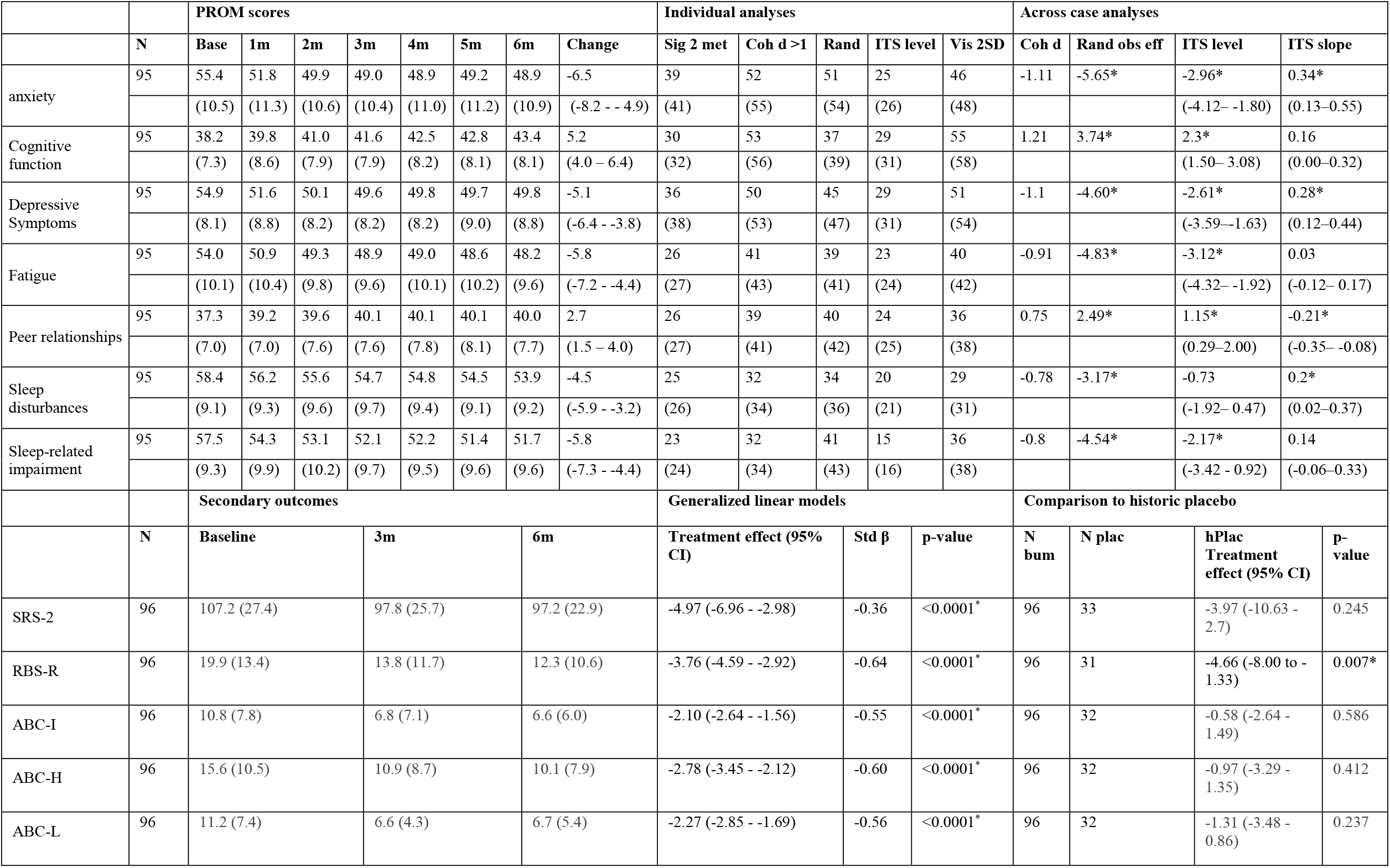
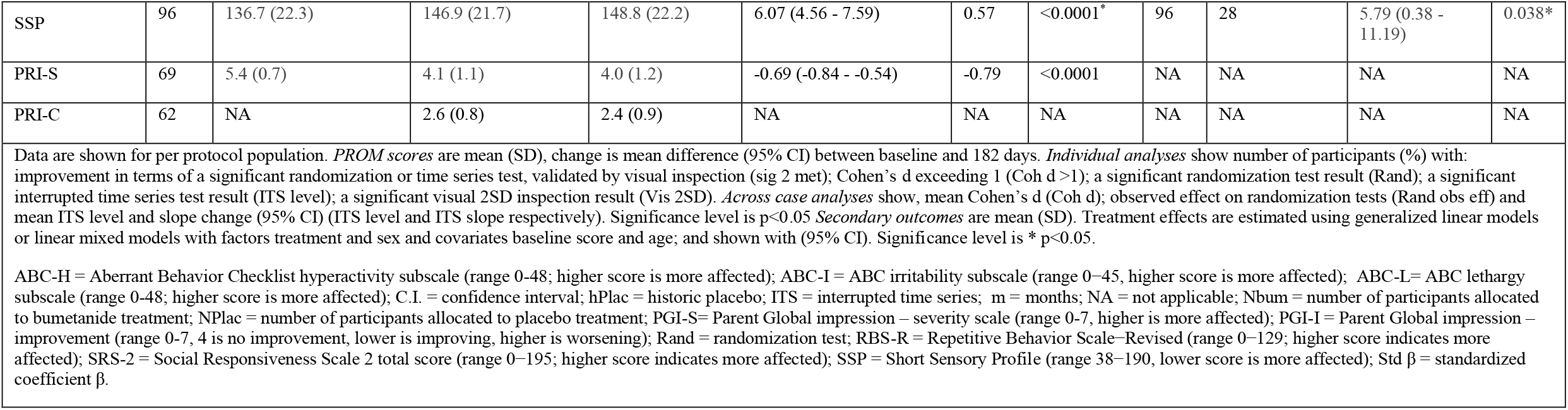
Changes in Primary and secondary outcome measures.

The most frequent improvements were observed for anxiety and depressive symptoms, affecting 41% and 38% of participants, respectively. The largest individual effect sizes were seen for cognitive function and anxiety, with Cohen’s *d*> 1 in 56% and 55% of participants, respectively. Patterns of co-occurring improvement most commonly involved anxiety in combination with depressive symptoms, fatigue, or sleep disturbance (Figure 2E). To evaluate consistency across analytic approaches, we compared visual 2-SD inspection with statistical decision rules based on interrupted time series and randomization tests. Agreement between visual inspection and interrupted time series analyses was higher (88%) than with randomization tests (61%) (Tables S4–S5). No participants showed statistically significant worsening on any of the seven PROM domains during the study period.

### Secondary outcome measures

All conventional questionnaires improved at group-level (Table 2; Table S6-S7). Age significantly affected model fit in ABC-I, ABC-H and RBS, sex did not. No correlation was found between bumetanide dose and outcome change. Superiority analyses versus historical placebo showed significant effects for RBS-R (SMD -4.66, 95%CI -8.00 to -1.33, *p*=.007; Figure S3), and SSP (SMD 5.79, 95%CI 0.38 to 11.19, *p*=.0376). This seemed consistent with the earlier effects found in the RBS-R and indicated an improvement on direct measurements of atypical sensory reactivity. Change in SSP correlated with change in anxiety and depressive symptoms (Pearson r>0.4) (Figure S4). No evidence of selection bias was observed between prior and new cohorts (*p*>.05, Figure S3).

### Adverse events

Adverse events (Table 3) were mild to moderate and resolved. One serious adverse event occurred: hip fracture after a sports incident and was probably unrelated to bumetanide treatment. Common expected adverse events included orthostatic complaints (37%), hypokalemia (36%), increased diuresis (35%), headache (20%) and dehydration (15%) and typically improved after increased fluid intake. In contrast with earlier studies, 17% of participants experienced dyspepsia (presented as decreased appetite), which fluctuated during the course of bumetanide treatment and showed no relation with bumetanide dose adjustments, although often resolved by decreasing the potassium chloride dose. Potassium levels remained above 2.8mmol/L (Table S8).

**Table 3:**
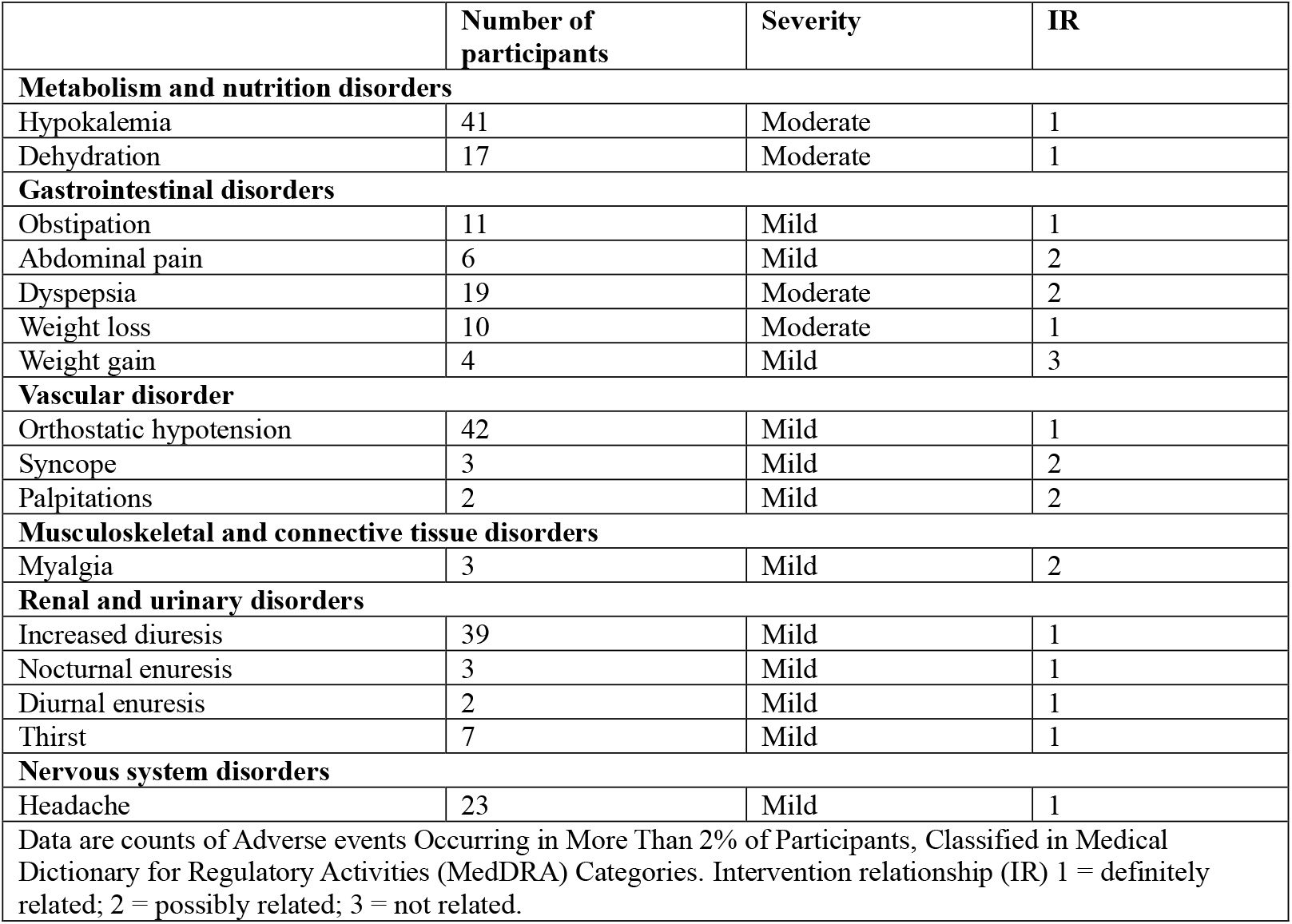
Adverse events.

## Discussion

Progress in pharmacological treatment development for NDDs has been limited not only by heterogeneous treatment responses, but also by a mismatch between targeted mechanisms, outcome measures, and trial designs. In this proof-of-concept study, we addressed two of these challenges by combining a parent-cocreated sensory reactivity PROMset with a SCED in a cohort enriched for atypical sensory processing. We demonstrate that daily health outcomes captured by this PROMset are (i) substantially impaired relative to reference populations, (ii) closely related to sensory reactivity severity, and (iii) sensitive to within-individual change over time. High interest and completion rates further support the feasibility of SCED methodology and parent-proxy reporting. Together, these findings indicate that combining sensory reactivity–focused PROMs with SCEDs enables detection of clinically meaningful, individualized treatment effects that may be obscured in conventional designs.

Using bumetanide as an example intervention, we demonstrate that the PROMset is sensitive to within-individual change over time. After six months, 83% of participants showed improvement on at least one PROMIS item bank, with approximately half improving across multiple domains, indicating that changes were often multidimensional rather than confined to a single symptom domain. At the level of individual domains, 24-41% showed clinically meaningful and statistically significant improvements, highlighting the limitation of single fixed primary outcomes in heterogenous populations: If a single outcome had been used, treatment-related changes might have been attenuated or missed. In contrast, the SCED framework enables detection of these individualized response patterns by focusing on within-person trajectories.

Prior PROM research on minimal clinically important differences indicates that changes of 3– 7 T-score points are meaningful to patients and their proxies (Schuchard et al., 2022; Selewski et al., 2017; Thissen et al., 2016; Tulsky et al., 2019). This range is substantially lower than the baseline differences observed between our cohort and reference populations (Table S1) and closely aligns with the mean within-person changes observed after six months (Table 2). Together, this supports the interpretation that PROMIS item banks are sufficiently sensitive to detect clinically relevant change in this population. At the same time, the proportion of individuals showing improvement on at least one PROMIS domain likely exceeds the proportion of true clinical responders. Future work should therefore focus on establishing robust response criteria that distinguish specific treatment effects from nonspecific or placebo-related changes, potentially by integrating multimodal measures such as electrophysiological biomarkers alongside patient-reported outcomes.

Despite inclusion of multiple NDDs and enrichment for atypical sensory reactivity, baseline scores on conventional measures were comparable to unselected ASD cohorts (Charman et al., 2017). In addition, PROMset impairments are in line with earlier reports in autism cohorts for Anxiety ((Schuchard et al., 2022), Fatigue (Schuchard et al., 2022), Sleep disturbance (Meltzer, Forrest, de la Motte, & Bevans, 2020; Schuchard et al., 2022), Sleep related impairments (Meltzer et al., 2020; Schuchard et al., 2022) and Peer relations (Toomey et al., 2016). Scores on the item banks Depressive symptoms and Cognitive function have not previously been evaluated in similar cohorts, although Depressive symptoms performed similar in an unspecified child and adolescent psychiatric cohort (Blomqvist, Chaplin, Henje, & Dennhag, 2025). These findings support generalizability to ASD populations.

The pattern of response may provide insight into potential underlying therapeutic mechanisms. We hypothesize that interventions such as bumetanide, may reduce anxiety and mood instability by improving sensory processing through modulation of the excitation-inhibition (E/I) balance, a framework that describes how neural systems regulate appropriate responses to sensory input (Ben-Ari, 2017; Henderson et al., 2012; Qin et al., 2015). Several phase-II bumetanide trials have already shown bumetanide’s effects on proxy measures of E/I balance alongside clinical improvements in responsive subsets (Juarez-Martinez et al., 2023; Juarez-Martinez et al., 2022; Lemonnier et al., 2012; Lemonnier et al., 2017; Sprengers et al., 2021; van Andel et al., 2022; van Andel, Sprengers, Konigs, de Jonge, & Bruining, 2024; van Andel et al., 2020; Zhang et al., 2020). This misalignment of outcome measures is particularly evident in the recent phase-III registration trial of bumetanide, which did not demonstrate effects on core symptoms in a broad population (Fuentes et al., 2023). However, exit interviews from that study reported improvements in domains such as serenity, cognitive function, sleep, and emotional well-being (Hawken et al., 2024). These findings suggest that treatment-related changes may occur in domains not captured by conventional outcome measures. In this context, the present results indicate that the PROMset is sensitive to changes in mood, cognition, and sleep that may otherwise remain undetected in traditional trial designs.

To ensure that observed changes reflect structured treatment dynamics rather than random fluctuation or bias, we incorporated several layers of methodological rigor. The multiple-baseline design allowed within-subject variability to serve as a control, while interrupted time series analyses and randomization testing enabled assumption-free statistical inference. The longitudinal structure of the SCED allowed us to pinpoint when changes emerged, confirming they followed the start of the intervention rather than during baseline. At the group level, comparisons with historic placebo data provided additional context. We also addressed potential responder bias inherent to post-trial access designs. We anticipated that the cohort might be enriched for individuals with prior favorable responses to bumetanide or for participants previously allocated to placebo. Contrary to these expectations, only one third of individuals anecdotally or questionnaire-defined as potential responders in previous trials enrolled in the study. Moreover, participants with prior bumetanide exposure did not differ in treatment response from those without prior exposure, nor from newly enrolled, treatment-naïve participants. These findings suggest that the cohort was not selectively enriched for bumetanide responders. Importantly, reasons for non-participation did not differ between individuals previously allocated to bumetanide or placebo. Participation appeared primarily driven by practical factors, including the number of required visits, travel distance, and venipuncture procedures, rather than treatment expectations. Lastly, we should note that part of the study was conducted during the COVID-19 pandemic, for which restrictions ended on March 22, 2022, in the Netherlands. Forty-five participants provided informed consent during the pandemic, of whom 22 completed the study before pandemic-related restrictions were lifted. Comparisons revealed no differences in outcomes between participants who completed the study during versus after the pandemic, indicating that pandemic-related factors are unlikely to have materially influenced the results. Collectively, these safeguards strengthen confidence in the internal validity and robustness of the observed patterns of change.

Despite these strenths, we acknowledge that the absence of masking and a concurrent placebo condition limits causal attribution of observed changes to bumetanide, as effects may partly reflect spontaneous improvement, placebo effects, regression to the mean, or effects from non-specific influences. Notably, the primary aim of this study was not to establish definitive efficacy, but to evaluate whether a clinically relevant PROM-based framework combined with a SCED can detect individualized treatment dynamics. From this perspective, the findings support the feasibility and sensitivity of this approach for capturing within-person change over time. Future studies incorporating placebo-controlled designs will be essential to establish causal treatment effects and to refine response criteria across outcome domains, thereby determining the role of this framework in drug development for heterogeneous NDD populations.

## Conclusion

This proof-of-concept study demonstrates that combining a parent-cocreated relevant sensory reactivity PROMset with a SCED trial, allows for the detection of meaningful change at the individual level, including in domains that are poorly captured by conventional symptom-based outcomes. These findings align with our broader synthesis that translational challenges in NDD treatment development often reflect misalignment between mechanisms, populations, and outcome measures rather than absence of biological effects. Together, this framework provides a practical and scalable approach to inform outcome selection, trial design, and regulatory evaluation. Ultimately, it supports a shift toward outcome strategies that align clinical research more closely with the lived experiences and priorities of children with NDDs and their families.

## Ethics statement

The study was conducted in accordance with the provisions of the Declaration of Helsinki and Good Clinical Practice. This protocol was approved by the medical ethical review board of UMC Utrecht (METC Utrecht) on 6/10/2020 (protocol number 20-356/G-X-M). Study safety was overseen by the monitoring board of Amsterdam. All participants or their legal representatives signed informed consent and received no financial compensation. This study was registered with the EudraCT trial registry (EudraCT 2020–002196-35).

## Author Contributions

L. Geertjens*: conceptualisation, data curation, formal analysis, investigation, methodology, project administration, software, supervision, validation, visualisation, writing – original draft, and writing – review & editing.

G. Cristian*: conceptualisation, formal analysis, methodology, software, validation, visualisation, writing – original draft, and writing – review & editing.

J.R. Ramautar: conceptualisation, investigation, supervision, and writing – review & editing.

L. Haverman: software, validation, and writing – review & editing.

B.D. Schalet: software, validation, and writing – review & editing.

K. Linkenkaer-Hansen: conceptualisation, funding acquisition, supervision, visualisation, and writing– review & editing.

G.J van der Wilt: conceptualisation, funding acquisition, methodology, software, supervision, validation, writing – original draft, and writing – review & editing.

J.J Sprengers*: conceptualisation, data curation, methodology, project administration, supervision, validation, visualisation, writing – original draft, and writing – review & editing.

H. Bruining*: conceptualisation, funding acquisition, methodology, project administration, resources, supervision, validation, visualisation, writing – original draft, and writing– review & editing.

*These authors contributed equally.

## Data availability

The data that support the findings of this study are available upon request from the corresponding author. These data are not publicly available due to privacy or ethical restrictions.

