## Supplementary information for "Assessing Parent-cocreated Sensory Reactivity Outcomes in Children with Neurodevelopmental Disorders Undergoing Bumetanide Treatment: A Multiple-Baseline Single-Case Experimental Design"

**Appendix S1: statistical analyses**

Visual-2SD-inspection (Horner & Swoboda, 2014; Parker & Brossart, 2003): Longitudinal PROMset scores were graphed as a time-series and smoothed using a lag-2 moving average (Figure 2A). A 2SD band from the baseline mean was visualized, unless a significant baseline trend was found in which case a 1SD-band was used (*n*=16-28; table S1) (Manolov R, 2017). A significant individual response was defined by two or more consecutive treatment phase points outside the 2SD band (figure 2A), or ≥25% of points outside the 1SD band, confirmed by two raters (LG, GC) (Hoogeboom et al., 2012).

Interrupted time series (ITS) analysis: ITS is particularly valuable for evaluating the impact of interventions when randomization is not feasible, as it allows for the identification of trends and shifts in data that occur before and after an intervention. Three key components were evaluated: change in level (immediate mean shift), mean difference (effect size, Cohen's d), and slope change (rate of change in score over time) between phases. Baseline trends were modeled as covariates to control for instability. At the group level, hierarchical models incorporated subject-specific random slopes to account for individual variability. Individual effect sizes were averaged to estimate group-level effects.This approach allows for the incorporation of variability among individual responses while providing a means to estimate the group-level effects.

Randomization test: This non-parametric test evaluates the mean difference between baseline and treatment. In the calculation, the data is randomly shuffled between the baseline and intervention phases. For each permutation of the dataset, the mean difference is recalculated. This procedure is repeated a number of times equal to the total number of possible permutations, thereby generating a permutation distribution of mean differences under the null hypothesis, which posits that there is no effect of the intervention. The null hypothesis is rejected when the observed mean difference falls within the extreme tail (top 5% values) of the permutation distribution.

**Table S1: Number of cases per PROM with a significant baseline trend**

| Anxiety | Cognitive function | Depressive symptoms | Fatigue | Peer Relationships | Sleep disturbance | Sleep-related impairment |
|---|---|---|---|---|---|---|
| 21 | 28 | 19 | 18 | 16 | 26 | 26 |
| Per protocol population (n=95); Data are n. | | | | | | |

**Table S2: PROM scores in the intention to treat population compared to the reference population**

| PROM questionnaire | Research population | Reference population | Difference | p-value |
|---|---|---|---|---|
| Anxiety | 55.5 (10.4) | 43.0 (11.6) | 12.5 (10.5 – 14.4) | <0.001 |
| Cognitive function | 38.4 (7.1) | 50.0 (10.0) | -11.6 (-13.0 – -10.3) | <0.001 |
| Depressive symptoms | 55.3 (8.0) | 43.5 (10.0) | 11.8 (10.3 -13.3) | <0.001 |
| Fatigue | 54.4 (9.7) | 39.6 (11.3) | 14.7 (12.9 -16.5) | <0.001 |
| Peer relationships | 37.2 (6.8) | 50.2 (10.4) | -13.0 (-14.2 - -11.7) | <0.001 |
| Sleep disturbance[1] | 59.3 (53.5 - 65.8) | 50.0 (8.9) | 9.2 | <0.001 |
| Sleep-related impairment[1] | 59.3 (53.3 - 63.7) | 50.1 (8.9) | 9.2 | <0.001 |

Intention to treat population (n=113). Data are mean (SD), and mean change (95% CI), or n (%). [1]Data with non-normal distribution. Data are shown as median (IQR) and median difference. PROM = patient reported outcome measure (higher scores indicate the construct is more present).

**Figure S1: Correlation between baseline PROMIS scores and SSP**

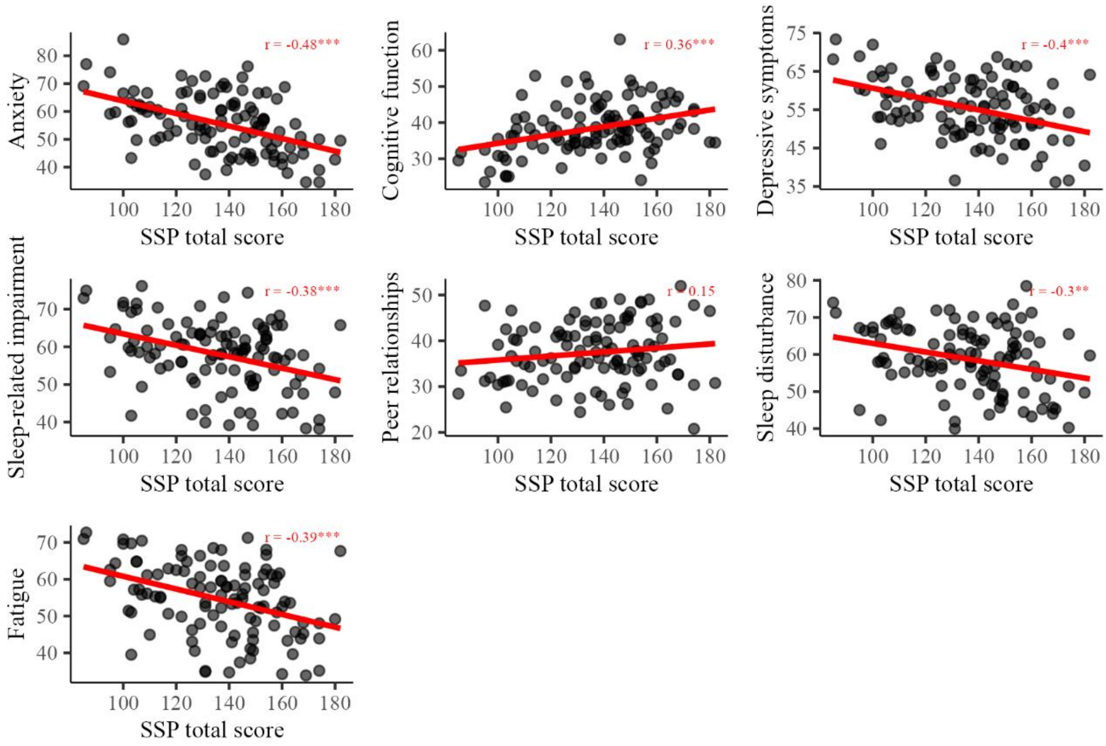


PROM = patient reported outcome measure (0-100, higher score indicates that the measured construct is more present); SSP = Short Sensory Profile (range 38−185, lower score is more affected).

**Table S3: Mean and SD for PROMs, split in previous and new cohort (per protocol population, LOCF)**

See separate file because of the size of the table

**Figure S2: PROMIS change over time**

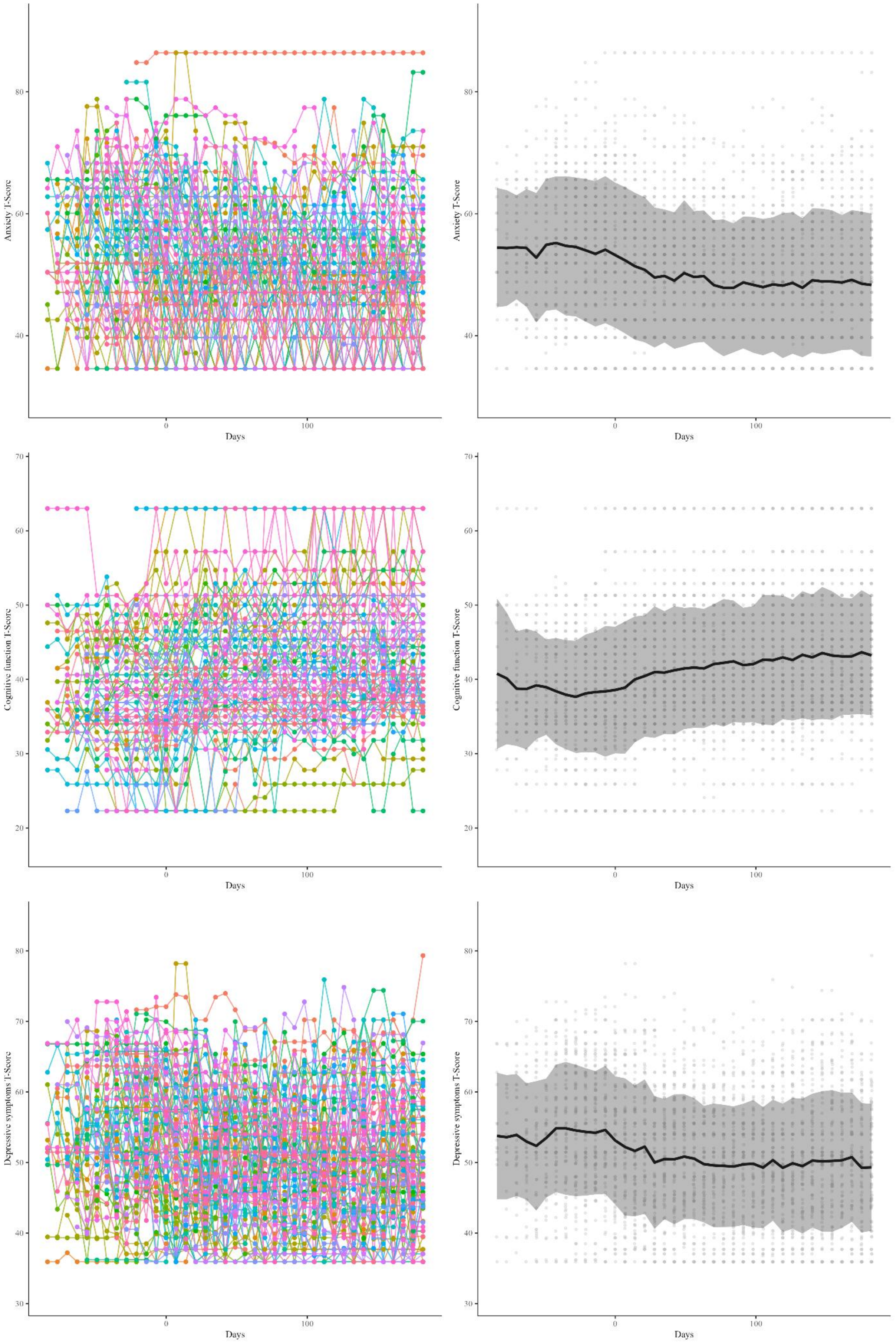

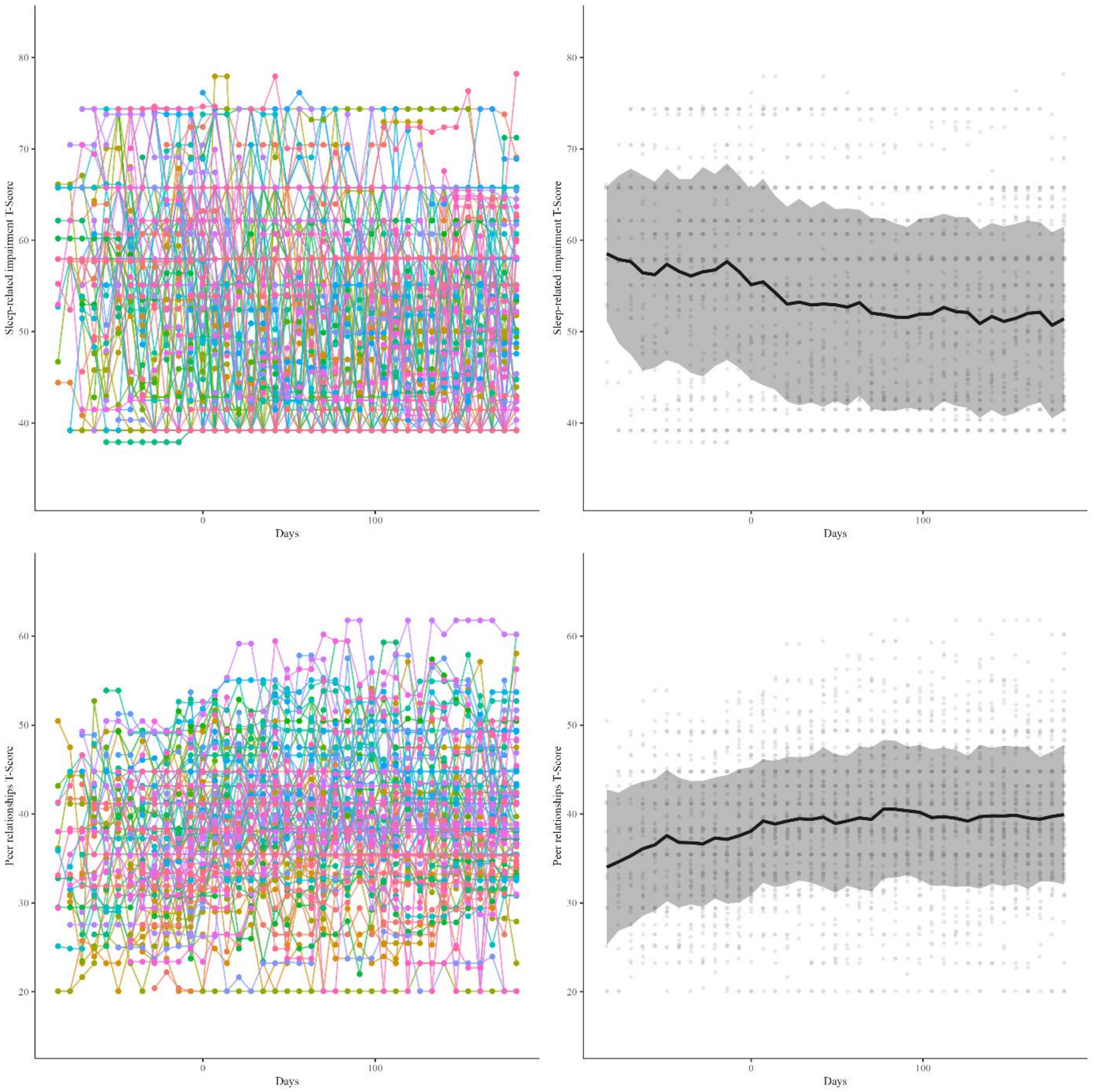
Sleep-related impairment T-Score
80
70
60
50
40
0
100
Days
Sleep-related impairment T-Score
80
70
60
50
40
0
100
Days
Peer relationships T-Score
60
50
40
30
20
0
100
Days
Peer relationships T-Score
60
50
40
30
20
0
100
Days

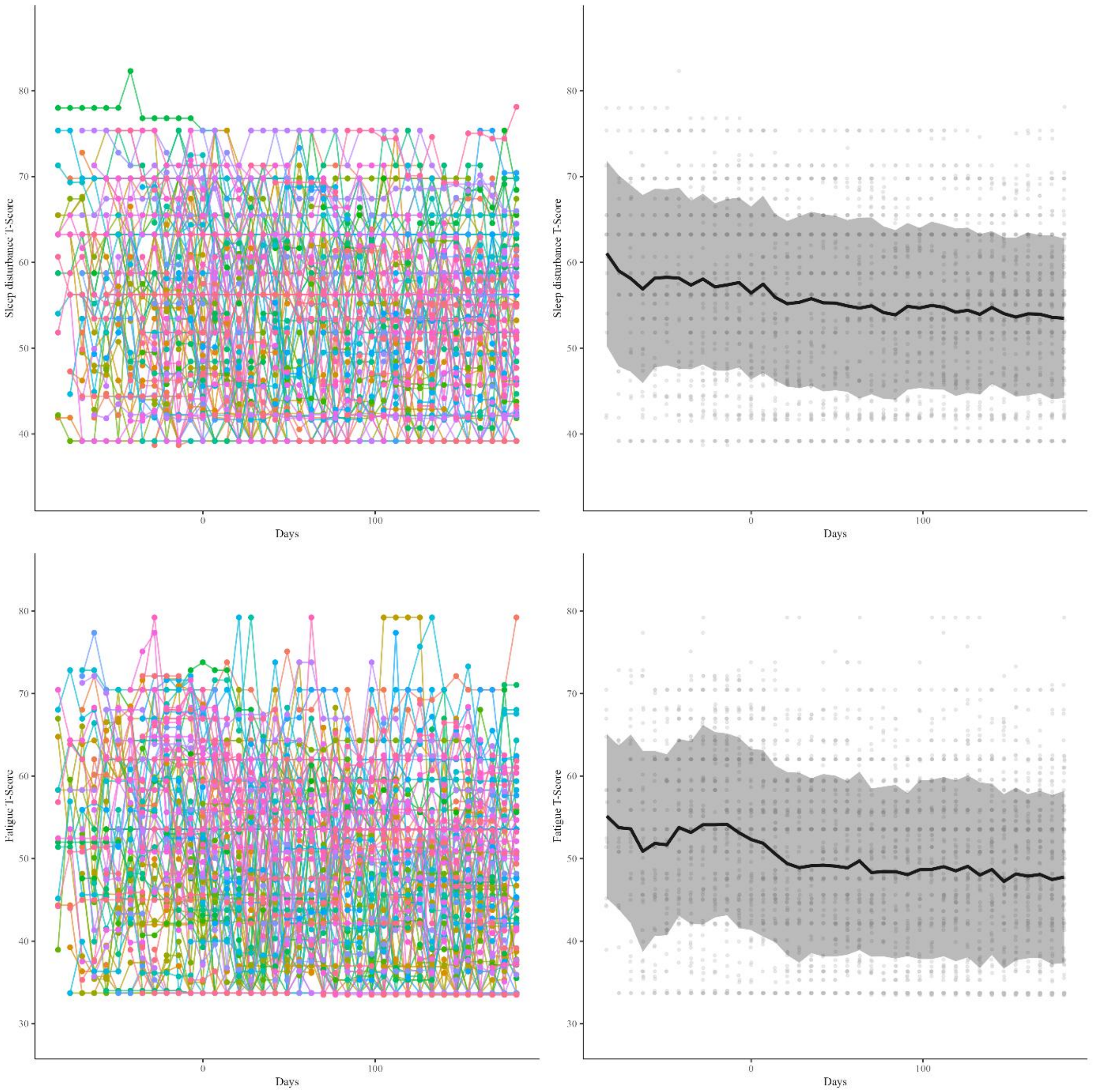


Left panel: Individual PROM score over time for the per protocol population (n=95) with last observation carried forward.; Right panel: mean (±1 SD) PROM score over time.

PROM = Patient reported outcome measure (Higher scores indicate the construct is more present).

### Table S4: Visual validation of significant results per statistical analysis

| Test | Anxiety | Cognitive function | Depressive symptoms | Fatigue | Peer Relationships | Sleep disturbance | Sleep-related impairment | Average |
|---|---|---|---|---|---|---|---|---|
| Randomization | 64 (67%) | 62 (65%) | 64 (67%) | 53 (56%) | 50 (53%) | 59 (62%) | 51 (54%) | 58 (61%) |
| Time series | 95 (100%) | 72 (76%) | 79 (83%) | 83 (87%) | 71 (75%) | 90 (95%) | 95 (100%) | 84 (88%) |
| Per protocol population (n=95); Data are n (%) | | | | | | | | |

**Table S5: Individual outcome for all three statistical tests per PROM**

See separate file because of the size of the table

### Table S6: subscales of conventional questionnaires (ITT)

| Subscale | Baseline score |
|---|---|
| **SRS-2** | |
| awareness | 10.53 (3.31) |
| cognition | 19.26 (5.69) |
| communication | 35.34 (10.87) |
| motivation | 18.69 (6.61) |
| preoccupation | 23.27 (7.71) |
| **RBS-R** | |
| compulsive | 2.73 (2.83) |
| restricted | 1.88 (1.83) |
| ritualistic | 4.13 (3.24) |
| sameness | 6.69 (5.42) |
| selfinjurious | 1.27 (2.21) |
| **SSP** | |
| auditory filtering | 15.89 (4.49) |
| movement sensitivity | 12.52 (2.96) |
| tactile sensitivity | 27.46 (5.76) |
| taste/Smell sensitivity | 16.03 (4.59) |
| underresponsive/seeks sensation | 25.91 (6.16) |
| visual/auditory sensitivity | 15.87 (4.80) |
| **ABC** | |
| hyperactivity | 15.90 (10.37) |
| irritability | 11.21 (7.67) |
| lethargy | 11.06 (7.24) |
| speech | 3.37 (2.88) |
| stereotypy | 4.31 (4.35) |
| Data are shown for the intention to treat population (n=113). Data are mean raw score (SD). ABC = Aberrant Behavior Checklist (higher score is more affected); RBS-R= Repetitive Behavior Scale–Revised (higher score indicates more affected); SRS-2= Social Responsiveness Scale 2 total score (higher score indicates more affected); SSP = Short Sensory Profile (lower score is more affected). | |

### Table S7: Mean and SD for conventional subscales, split in previous and new cohort

| | Per protocol population (n=96) | | | Previous cohort (n=27) | | | New cohort (n=69) | | |
|---|---|---|---|---|---|---|---|---|---|
| **Subscale** | **Baseline** | **3 months** | **6 months** | **Baseline** | **3 months** | **6 months** | **Baseline** | **3 months** | **6 months** |
| **SRS** | | | | | | | | | |
| awareness | 10.61 (3.34) | 9.79 (3.38) | 9.18 (3.46) | 10.74 (3.75) | 9.57 (3.46) | 9.22 (3.19) | 10.57 (3.19) | 9.86 (3.38) | 9.16 (3.59) |
| cognition | 19.09 (5.43) | 17.19 (5.19) | 17.08 (5.05) | 17.11 (5.41) | 16.33 (6.10) | 17.00 (4.79) | 19.87 (5.28) | 17.48 (4.87) | 17.12 (5.18) |
| communication | 35.42 (10.78) | 31.83 (9.87) | 31.75 (8.39) | 29.74 (10.56) | 28.95 (12.21) | 30.89 (8.21) | 37.64 (10.09) | 32.79 (8.87) | 32.09 (8.50) |
| motivation | 18.72 (6.82) | 17.04 (5.76) | 16.75 (5.65) | 15.22 (6.72) | 14.76 (7.29) | 15.78 (5.55) | 20.09 (6.40) | 17.79 (4.99) | 17.13 (5.69) |
| preoccupation | 23.32 (7.70) | 21.98 (7.43) | 22.47 (5.92) | 16.00 (7.06) | 16.29 (9.47) | 20.19 (6.52) | 26.19 (5.85) | 23.87 (5.51) | 23.36 (5.46) |
| **RBS** | | | | | | | | | |
| compulsive | 2.78 (2.77) | 1.89 (2.65) | 1.73 (2.26) | 2.89 (3.43) | 1.91 (3.62) | 1.78 (2.83) | 2.74 (2.48) | 1.89 (2.25) | 1.71 (2.02) |
| restricted | 1.97 (1.91) | 1.36 (1.56) | 1.21 (1.44) | 1.33 (1.18) | 0.77 (0.92) | 0.81 (1.18) | 2.22 (2.08) | 1.57 (1.68) | 1.36 (1.51) |
| ritualistic | 4.19 (3.20) | 3.33 (3.32) | 2.69 (2.93) | 3.63 (3.47) | 3.23 (3.66) | 2.33 (3.13) | 4.41 (3.08) | 3.37 (3.22) | 2.83 (2.86) |
| sameness | 6.77 (5.37) | 4.42 (4.19) | 4.00 (3.84) | 6.30 (6.09) | 3.91 (4.95) | 3.70 (5.49) | 6.96 (5.10) | 4.60 (3.92) | 4.12 (3.01) |
| selfinjurious | 1.19 (2.14) | 1.00 (2.21) | 0.95 (2.14) | 0.37 (0.84) | 0.23 (0.69) | 0.41 (1.01) | 1.51 (2.40) | 1.27 (2.49) | 1.16 (2.42) |
| **SSP** | | | | | | | | | |
| auditory filtering | 15.99 (4.69) | 18.50 (4.59) | 19.04 (4.54) | 17.67 (4.20) | 19.10 (4.32) | 20.30 (4.80) | 15.33 (4.74) | 18.30 (4.69) | 18.55 (4.37) |
| movement sensitivity | 12.45 (2.97) | 12.77 (2.67) | 12.59 (3.11) | 12.33 (3.04) | 12.76 (2.66) | 12.59 (2.86) | 12.49 (2.97) | 12.78 (2.69) | 12.59 (3.23) |
| tactile sensitivity | 27.50 (5.72) | 29.08 (4.95) | 29.23 (5.10) | 28.93 (6.49) | 29.52 (5.63) | 29.81 (5.82) | 26.94 (5.34) | 28.94 (4.74) | 29.00 (4.81) |
| taste/Smell sensitivity | 16.05 (4.61) | 16.39 (4.55) | 16.53 (4.48) | 16.78 (3.94) | 17.29 (4.26) | 17.26 (4.06) | 15.77 (4.84) | 16.10 (4.64) | 16.25 (4.63) |
| underresponsive/seeks sensation | 26.18 (6.21) | 27.94 (6.23) | 28.71 (5.58) | 27.89 (4.59) | 29.00 (4.74) | 30.33 (4.66) | 25.51 (6.65) | 27.59 (6.65) | 28.07 (5.81) |
| visual/auditory sensitivity | 15.93 (4.72) | 17.85 (4.86) | 18.56 (4.54) | 16.33 (5.02) | 18.24 (5.28) | 19.37 (5.56) | 15.77 (4.62) | 17.71 (4.75) | 18.25 (4.07) |
| **ABC** | | | | | | | | | |
| hyperactivity | 15.64 (10.52) | 10.89 (8.72) | 10.07 (7.94) | 13.26 (8.90) | 8.36 (7.44) | 8.89 (7.30) | 16.57 (11.01) | 11.78 (9.01) | 10.54 (8.18) |
| irritability | 10.82 (7.84) | 6.81 (7.06) | 6.62 (5.97) | 7.89 (6.40) | 4.73 (5.40) | 5.15 (5.50) | 11.97 (8.08) | 7.54 (7.45) | 7.20 (6.08) |
| lethargy | 11.23 (7.37) | 6.62 (4.28) | 6.69 (5.44) | 11.00 (8.47) | 5.64 (4.90) | 6.78 (6.59) | 11.32 (6.95) | 6.97 (4.03) | 6.65 (4.97) |
| inappropriate speech | 3.47 (2.97) | 2.42 (2.50) | 2.19 (2.38) | 3.30 (3.24) | 2.36 (2.40) | 2.04 (2.23) | 3.54 (2.88) | 2.44 (2.55) | 2.25 (2.45) |
| stereotypy | 4.45 (4.43) | 2.94 (4.09) | 2.60 (3.64) | 3.67 (4.64) | 2.41 (4.41) | 1.89 (3.19) | 4.75 (4.34) | 3.13 (3.99) | 2.88 (3.79) |

Per protocol population (n=95). Data are mean raw score (SD)
ABC = Aberrant Behavior Checklist (higher score is more affected); RBS-R = Repetitive Behavior Scale−Revised (higher score indicates more affected); SRS-2 = Social Responsiveness Scale 2 total score (higher score indicates more affected); SSP = Short Sensory Profile (lower score is more affected)

**Figure S3: Treatment effect on repetitive behavior**

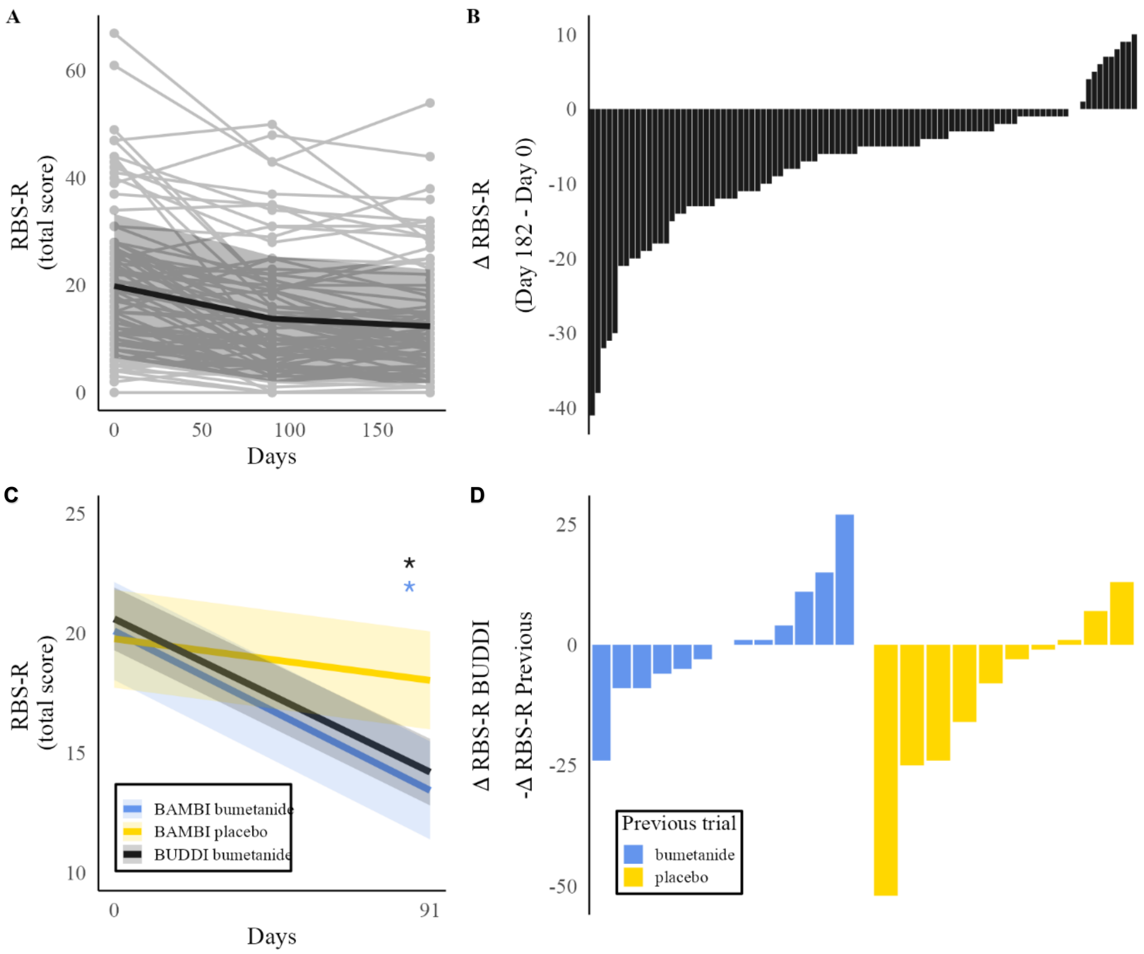


(A) Individual RBS-R total scores over time, including mean ±1SD band. (B) Individual RBS-R total score change between D182 and D0. A negative score indicates improvement; Each bar represents one participant. (C) Comparison of RBS-R scores in current trial to historic BAMBI trial, using linear mixed models. (D) Difference in individual RBS-R total score change between current trial and historic trial. A negative score indicated more improvement in the current trial; Each bar represents one participant. RBS-R= Repetitive Behavior Scale–Revised (range 0−129; higher score indicates more affected); Significance level * $p<0{\cdot}05$;

**Figure S4: Correlation between change PROM score and change in SSP total score**

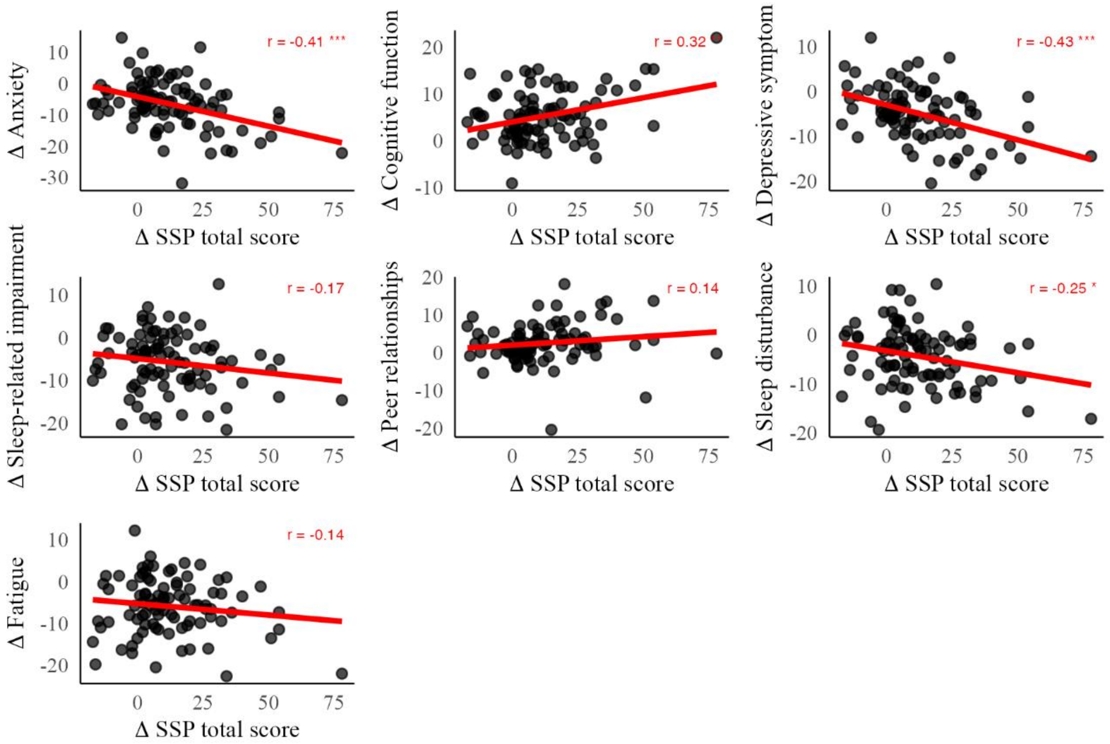


PROM = patient reported outcome measure (0-100, higher score indicates that the measured construct is more present); RBS-R = Repetitive Behavior Scale−Revised (range 0−129; higher score indicates more affected); SRS-2 = Social Responsiveness Scale 2 total score (range 0−195; higher score indicates more affected); SSP = Short Sensory Profile (range 38−185, lower score is more affected);

### Table S8: potassium values per participant per measurement

| | baseline | extra | d14 | extra | extra | d28 | extra | d56 | extra | extra | d91 | extra | extra | extra | extra | extra |
|---|---|---|---|---|---|---|---|---|---|---|---|---|---|---|---|---|
| 1 | 3.9 | | 3.4 | 3.6 | | 3.9 | 3.7 | 3.9 | | | 4.0 | | | | | |
| 2 | 4.4 | 3.9 | 4.3 | | | 3.7 | 4.1 | 3.9 | | | 3.9 | | | | | |
| 3 | 4.0 | | 3.8 | | | 3.9 | | 4.0 | 3.9 | | 3.8 | | | | | |
| 4 | 3.8 | | | | | 3.5 | | 3.8 | | | 3.8 | | | | | |
| 5 | 3.5 | | 3.4 | | | 3.6 | | 3.9 | | | 3.7 | | | | | |
| 6 | 4.4 | | 4.0 | | | 4.1 | | 3.8 | | | 3.8 | | | | | |
| 7 | 4.0 | | 4.1 | | | 3.5 | | 3.4 | | | 3.9 | | | | | |
| 8 | 3.9 | | 3.6 | | | 4.1 | | 4.1 | | | 3.7 | | | | | |
| 9 | 3.9 | | 4.3 | | | 3.7 | | 3.8 | | | 3.9 | | | | | |
| 10 | 4.1 | | 3.1 | 3.7 | | 4.3 | | 4.3 | | | 3.8 | | | | | |
| 11 | 3.6 | | 3.2 | 3.3 | | 3.3 | | 3.9 | | | 4.2 | | | | | |
| 12 | 4.3 | | 3.4 | 3.6 | | 3.3 | 4.0 | 3.8 | | | 3.5 | | | | | |
| 13 | 4.2 | | | | | | | 3.1 | 2.9 | 3.7 | NA | | | | | |
| 14 | 4.4 | | 3.3 | 3.7 | | NA | | 4.7 | | | 3.4 | 3.7 | | | | |
| 15 | 4.1 | | 2.9 | 3.2 | 4.1 | 3.9 | | 3.8 | | | 3.5 | | | | | |
| 16 | 4.1 | | 3.6 | | | 4.0 | | 4.0 | | | 3.9 | | | | | |
| 17 | 3.9 | | 3.9 | | | 3.9 | | 3.9 | | | 3.8 | | | | | |
| 18 | 4.1 | | 4.0 | | | 4.3 | | 4.4 | | | 4.2 | | | | | |
| 19 | 4.4 | | | | | 3.4 | 3.6 | 3.8 | | | 4.0 | | | | | |
| 20 | 4.1 | | | | | 3.2 | 4.1 | 3.5 | | | 3.6 | | | | | |
| 21 | 4.1 | | 3.4 | | | 4.1 | | NA | | | NA | | | | | |
| 22 | 4.0 | | 4.0 | | | 3.9 | | 3.9 | | | 4.1 | | | | | |
| 23 | 3.8 | | | | | 3.4 | 3.6 | 3.7 | | | 3.3 | 3.8 | 3.4 | | | |
| 24 | 4.0 | | 3.9 | | | 4.1 | | 3.9 | | | 3.9 | 3.9 | | | | |
| 25 | 4.5 | | | | | 3.9 | | | | | | | | | | |
| 26 | 4.5 | | 4.0 | | | 3.5 | | 3.9 | | | 3.7 | | | | | |
| 27 | 4.1 | | 3.3 | | | 4.2 | | 3.4 | | | 3.6 | 3.6 | | | | |
| 28 | 4.2 | | | | | 4.3 | | 4.4 | | | 5.3[*] | 3.8 | | | | |
| 29 | 3.8 | | 3.7 | | | 3.7 | | 3.8 | | | 3.7 | | | | | |
| 30 | 4.2 | | | | | 3.8 | | 3.9 | | | 3.5 | 3.7 | | | | |
| 31 | 4.1 | | 3.2 | 3.8 | | 3.7 | | 3.7 | | | 3.8 | | | | | |
| 32 | 4.2 | | 3.7 | | | 3.9 | | 4.1 | | | 3.8 | 3.7 | | | | |
| 33 | 3.9 | | 3.4 | | | 3.6 | | 3.8 | | | 3.6 | | | | | |
| 34 | 4.0 | | 3.4 | | | 3.7 | | 3.7 | | | 3.3 | 3.5 | | | | |
| 35 | 4.1 | | | | | 4.3 | | 3.7 | | | 3.8 | | | | | |
| 36 | 4.0 | | 4.7 | | | 4.2 | | 4.1 | | | 3.7 | | | | | |
| 37 | 3.9 | | | | | 3.9 | | 3.4 | | | 3.7 | | | | | |
| 38 | 4.2 | | | | | 3.2 | 3.5 | 3.5 | | | 3.5 | | | | | |
| 39 | 4.0 | | | | | 4.0 | | 3.7 | | | 3.3 | 3.4 | 3.5 | | | |

| | | | | | | | | | | | | | | | | |
|---|---|---|---|---|---|---|---|---|---|---|---|---|---|---|---|---|
| 40 | 4.4 | | 3.7 | | | 3.7 | | NA | | | NA | | | | | |
| 41 | 3.4 | | 2.9 | 3.6 | | 3.8 | | 3.8 | | | 3.6 | | | | | |
| 42 | 4.1 | | 3.5 | | | 4.3 | | 3.3 | 3.5 | | 3.3 | 3.9 | | | | |
| 43 | 4.4 | | 3.8 | | | 3.7 | | 3.7 | | | 3.6 | | | | | |
| 44 | 4.1 | | 3.9 | | | 3.7 | | 3.7 | | | 3.5 | | | | | |
| 45 | 4.6 | | 4.0 | | | 3.5 | | 4.5 | | | 3.8 | | | | | |
| 46 | 4.5 | | 4.2 | | | 4.1 | | 3.7 | | | 4.1 | | | | | |
| 47 | 4.5 | | 4.1 | | | 3.8 | | 4.0 | | | 3.8 | | | | | |
| 48 | 4.4 | | 3.6 | | | 4.4 | | 4.7 | | | 3.7 | | | | | |
| 49 | 4.1 | | 3.9 | | | 3.8 | | 3.2 | 3.3 | | 3.9 | | | | | |
| 50 | 4.2 | | 2.8 | 3.8 | | 5.0 | | 3.9 | | | 3.1 | 3.3 | 3.6 | | | |
| 51 | 3.9 | | F | | | 3.4 | | 3.7 | | | 3.5 | | | | | |
| 52 | 3.9 | | 4.0 | | | 3.5 | | 3.8 | | | 3.7 | | | | | |
| 53 | 3.8 | | 3.6 | | | 3.2 | 4.1 | 3.7 | | | 3.8 | | | | | |
| 54 | 4.3 | | 3.6 | | | 3.7 | | 3.3 | 3.6 | | 4.0 | 3.5 | 3.2 | 3.0 | 3.4 | 3.9 |
| 55 | 3.8 | | 4.2 | | | 4.0 | | 4.2 | | | 3.1 | 3.6 | 3.5 | | | |
| 56 | 4.3 | | 3.6 | | | 4.0 | | 3.8 | | | 4.0 | | | | | |
| 57 | 4.4 | | 4.3 | | | 3.7 | | 4.0 | | | 3.8 | | | | | |
| 58 | 3.9 | | 3.8 | | | 3.9 | | 3.7 | | | 3.8 | | | | | |
| 59 | 3.8 | | 3.5 | | | 3.8 | | NA | | | NA | | | | | |
| 60 | 3.9 | | 3.7 | | | 3.8 | | 3.5 | | | 3.7 | | | | | |
| 61 | 3.9 | | 3.8 | | | 3.4 | | 3.4 | | | 3.4 | | | | | |
| 62 | 3.9 | | 4.1 | | | 3.9 | | 3.8 | | | NA | | | | | |
| 63 | 4.3 | | 3.8 | | | 3.9 | | 3.8 | | | 3.7 | | | | | |
| 64 | 3.9 | | | | | 3.4 | | 3.5 | | | 3.7 | | | | | |
| 65 | 3.9 | | | | | 3.3 | | 3.7 | | | 3.6 | | | | | |
| 66 | 4.2 | | 5.1 | | | 3.3 | 3.5 | 3.7 | | | 3.3 | 3.8 | | | | |
| 67 | 4.3 | | 3.6 | | | 4.0 | | 3.9 | | | 3.7 | | | | | |
| 68 | 4.0 | | 3.7 | | | F | | 3.7 | | | NA | | | | | |
| 69 | 3.8 | | 3.5 | | | 3.9 | | 3.7 | | | 3.6 | | | | | |
| 70 | 4.1 | | 3.8 | | | 3.9 | | 3.8 | | | 3.9 | | | | | |
| 71 | 4.0 | | 3.6 | | | 3.4 | | 3.4 | | | 3.0 | 2.7 | 3.3 | 3.3 | 3.5 | 3.6 |
| 72 | 3.7 | | 4.2 | | | 3.8 | | 3.3 | | | 3.3 | 3.7 | 3.4 | | | |
| 73 | 4.6 | | 3.8 | | | 3.6 | | 3.4 | | | 3.5 | | | | | |
| 74 | 4.2 | | | | | 3.4 | | 3.5 | | | 3.6 | | | | | |
| 75 | 3.9 | | 3.3 | 3.3 | | 3.4 | | 3.4 | | | 3.7 | | | | | |
| 76 | 4.3 | | | | | 4.0 | | 3.6 | | | 3.8 | | | | | |
| 77 | 4.2 | | 3.1 | 3.1 | | 3.6 | | 3.5 | | | 3.7 | | | | | |
| 78 | 4.4 | | 4.1 | | | 4.0 | | 4.0 | | | 3.9 | | | | | |
| 79 | 3.9 | | 3.6 | | | 3.5 | | 3.5 | | | 3.5 | | | | | |
| 80 | 3.8 | | 3.1 | | | 3.8 | | 3.9 | | | 3.3 | 3.8 | 3.7 | | | |
| 81 | 4.1 | | 3.8 | | | 3.8 | | 3.6 | | | 3.5 | | | | | |
| 82 | 3.8 | | 3.7 | | | 3.5 | | 3.8 | | | 3.8 | | | | | |

| | | | | | | | | | | | | | | | | |
|---|---|---|---|---|---|---|---|---|---|---|---|---|---|---|---|---|
| 83 | 4.0 | | | | | NA | | 3.0 | | | 3.8 | | | | | |
| 84 | 3.9 | | 4.0 | | | 3.6 | | 3.7 | | | 3.8 | | | | | |
| 85 | 4.6 | | | | | 4.0 | | 3.2 | 3.7 | | 4.1 | | | | | |
| 86 | 3.8 | | 3.6 | | | 3.8 | | 3.7 | | | 3.2 | 3.4 | 3.7 | | | |
| 87 | 4.0 | | 3.4 | | | 4.2 | | 3.3 | F | 3.0 | 3.5 | | | | | |
| 88 | 4.4 | | 4.3 | | | 4.1 | | 4.0 | | | 4.1 | | | | | |
| 89 | 4.3 | | NA | | | 3.8 | | 3.6 | | | 3.8 | | | | | |
| 90 | 4.6 | | 4.5 | | | 4.4 | | 3.6 | | | 4.0 | | | | | |
| 91 | 4.4 | | 3.9 | | | 3.6 | | 3.7 | | | 3.7 | | | | | |
| 92 | 4.0 | | 4.0 | | | 4.5 | | 4.1 | | | 4.0 | | | | | |
| 93 | 4.1 | | 3.7 | | | 3.4 | | 3.8 | | | 3.7 | | | | | |
| 94 | 3.9 | | | | | 3.8 | | 3.5 | | | 3.8 | | | | | |
| 95 | 4.3 | | 4.0 | | | 3.9 | | 4.1 | | | 4.4 | | | | | |
| 96 | 4.0 | | 3.2 | 3.7 | | 3.3 | 3.7 | 3.4 | | | 3.4 | | | | | |
| 97 | 3.9 | | 3.8 | | | 3.5 | | 3.7 | | | 4.1 | | | | | |
| 98 | 4.3 | | 3.4 | | | 4.1 | | 3.8 | | | 3.8 | | | | | |
| 99 | 4.1 | | 3.4 | | | 3.9 | | 3.8 | | | 3.6 | | | | | |
| 100 | 4.4 | | 3.6 | | | 3.6 | | 3.9 | | | 3.7 | | | | | |
| 101 | 3.7 | | NA | | | NA | | NA | | | NA | | | | | |
| 102 | 3.8 | | 4.2 | | | 3.4 | | 3.3 | 3.1 | 3.1 | 3.0 | 2.8 | 3.2 | 4.0 | 3.4 | 3.4 |
| 103 | 3.8 | | | | | 3.8 | | 3.3 | 3.8 | | 3.8 | | | | | |
| 104 | 3.7 | | 4.1 | | | 3.9 | | 3.2 | 3.3 | | 3.8 | | | | | |
| 105 | 3.5 | | 3.6 | | | 3.5 | | 3.5 | | | 3.2 | 3.3 | 3.4 | | | |
| 106 | 4.1 | | 3.8 | | | 3.7 | | 4.1 | | | 4.0 | | | | | |
| 107 | 3.9 | | NA | | | 3.7 | | 3.5 | | | 3.4 | | | | | |
| 108 | 3.9 | | 3.5 | | | 4.2 | | 4.0 | | | 3.6 | | | | | |
| 109 | 4.2 | | 4.2 | | | 3.3 | 3.5 | 3.5 | | | 3.6 | | | | | |
| 110 | 3.8 | | 3.5 | | | 3.1 | 3.6 | 3.3 | 3.3 | | 3.4 | | | | | |
| 111 | 4.0 | | 3.7 | | | 3.8 | | 3.7 | | | 3.8 | | | | | |
| 112 | 4.0 | | 3.5 | | | 3.9 | | 4.0 | | | 4.0 | | | | | |
| 113 | 4.1 | | 3.6 | | | 3.7 | | 3.9 | | | 4.8 | | | | | |

Data are mmol/L;
*= haemolytic sample; NA = not applicable
